# Prospective phenotyping of CHAMP1 disorder indicates that coding mutations do not act through haploinsufficiency

**DOI:** 10.1101/2023.04.17.23288527

**Authors:** Tess Levy, Thariana Pichardo, Hailey Silver, Bonnie Lerman, Jessica Zweifach, Danielle Halpern, Paige M Siper, Alexander Kolevzon, Joseph D Buxbaum

## Abstract

CHAMP1 disorder is a genetic neurodevelopmental condition caused by mutations in the CHAMP1 gene that result in premature termination codons. The disorder is associated with intellectual disability, medical comorbidities, and dysmorphic features. Deletions of the *CHAMP1* gene, as part of 13q3 deletion syndrome, have been briefly described with the suggestion of a milder clinical phenotype. To date, no studies have directly assessed differences between individuals with mutations in *CHAMP1* to those with deletions of the gene. We completed prospective clinical evaluations of 16 individuals with mutations and eight with deletions in *CHAMP1*. Analyses revealed significantly lower adaptive functioning across all domains assessed (i.e., communication, daily living skills, socialization, and motor skills) in the mutation group. Developmental milestones and medical features further showed difference between groups. The phenotypes associated with mutations, as compared to deletions, indicate likely difference in pathogenesis between groups, where deletions are acting through CHAMP1 haploinsufficiency and mutations are acting through dominant negative or gain of function mechanisms, leading to a more severe clinical phenotype. Understanding this pathogenesis is important to the future of novel therapies for CHAMP1 disorder and illustrates that mechanistic understanding of mutations must be carefully considered prior to treatment development.

## Introduction

*CHAMP1*, or chromosome alignment maintaining phosphoprotein 1, is a gene located on the long arm of chromosome 13 and mutations in the gene are associated with CHAMP1-related neurodevelopmental disorder, or CHAMP1 disorder (Garrity 2021; Hempel 2015; Isidor 2016; Itoh 2011; Levy 2022b; Tanaka 2016). There are only a modest number of published reports on CHAMP1 disorder, but analysis of over 1000 individuals serially ascertained for intellectual or developmental delay (IDD) indicate that as many as 1 in 500 individuals with IDD may carry mutations in this gene (Disorders. 2015), indicating profound under-diagnosis of this disorder. Published cases all presented with global developmental delays (speech, motor) and intellectual disability (ID). A majority had hypotonia, gait abnormalities, ocular abnormalities, gastrointestinal abnormalities (including cyclical vomiting), microcephaly, and visual abnormalities. Recurrent infections are reported as well. Behavioral findings included anxiety, moderate to severe attention-deficit/hyperactivity disorder, autism spectrum disorder traits, and sensory reactivity abnormalities.

CHAMP1 is a zinc finger (ZNF) phosphoprotein that was first described as being required for maintenance of kinetochore-microtubule attachment, which is critical for accurate chromosome segregation (Itoh 2011). CHAMP1 contains C2H2-type zinc finger domains in the N- and C-terminal regions, which flank a core region that includes multiple SPE (PxxSPExxK), WK (SPxxWKxxP), and FPE (FPExxK) motifs. CHAMP1 is part of the heterochromatin protein 1 (HP1) complex, which is involved in many nuclear processes including heterochromatin formation and gene silencing, transcriptional activation or arrest, regulation of binding of cohesion complexes to centromeres, and DNA repair (van Wijnen 2021). This complex regulates the expression of cell and tissue-type specific genes during development, including in neuronal maturation during cortical development (Oshiro 2015). Several other IDD genes (e.g., ADNP, CHD4, CTCF, EHMT1, and POGZ) are found in HP1 complexes (Mosch 2011; Ostapcuk 2018), underscoring the importance of this complex in CNS development. The C-terminal ZNF domain of CHAMP1 mediates the interaction of CHAMP1 with both HP1 and POGZ, MAD2L2 binds to CHAMP1 through interaction with the WK-motif-rich middle region of the protein, and the N- and C-terminal regions are responsible for chromosome and spindle localization (Isidor 2016; Itoh 2011).

Reported pathogenic CHAMP1 sequence variants have all resulted in premature termination codon (PTC) mutations, either nonsense, frameshift, or splice site variants; to date, no missense variants in *CHAMP1* have been clinically reported as pathogenic. When biological parents were available, all cases were confirmed de novo. Several recurrent variants have been noted, specifically Arg398* and Arg497*. In genetic disorders, when only PTC mutations are seen as causative of disease, it is frequently assumed that the mechanism is a loss of function (LoF) leading to haploinsufficiency. However, CHAMP1 is expressed from the terminal exon of the gene and hence the resultant RNA is predicted to escape nonsense mediated decay (NMD). This raises the possibility that truncated proteins are expressed that may function through dominant negative or gain of function (GoF) mechanisms. While this has been speculated on in the past, it has not been conclusively resolved; currently, as the possibility of gene therapy becomes more feasible and with several groups considering gene therapy in CHAMP1 disorder, it is urgent to better understand the biological mechanisms of clinical CHAMP1 mutations.

Evidence for a LoF mechanism can be garnered by examining the phenotypes of individuals carrying a deletion over the gene of interest. For example, both PTCs in *SHANK3* and deletions encompassing *SHANK3* produce overlapping phenotypes, consistent with a LoF mechanism (De Rubeis 2018; Fu 2022; Levy 2022a). In the current report, we carried out targeted phenotyping of individuals with 13q3 deletions that included CHAMP1 or with CHAMP1 mutations, using methods that we have reported for individuals with CHAMP1 mutations (Levy 2022b). This allowed us to directly compare phenotypes in individuals with 13q3 deletions with those with CHAMP1 mutations to determine whether the results are consistent with CHAMP1 PTC mutations operating through a LoF mechanism.

## Methods

### Participants

Participants ranged from 1.4 to 29.8 years of age (8.79 ± 6.7) and included 12 females and 12 males (Table 1). Participants were grouped into CHAMP1 mutation group and 13q3 deletion group. Participants in the mutation group had a sequence variant within *CHAMP1* classified as pathogenic or likely pathogenic according to the American College of Medical Genetics and Genomics and Association for Molecular Pathology guidelines. Participants in the deletion group had a microdeletion on chromosome 13 which included *CHAMP1* (Figure 1). All genetic reports were reviewed by a genetic counselor. Two additional participants with ring chromosome 13 were recruited and reported here, but not included in either grouping, due to both genetic and clinical differences.

**Table 1.** Participant Demographics.

| Table 1. Participant Demographics |  |  |  |
| --- | --- | --- | --- |
|  | Mutations | Deletions | All |
| N | 16 | 8 | 24 |
| Age (years) | 1 - 29 (M=9.4; SD=7.7) | 1 – 13 (M=7.6; SD=4.1) | M=8.79; SD=6.7 |
| Sex (Female: Male) | 9:7 | 3:5 | 12:12 |
| Ethnicity | 1/16 Hispanic / Latinx | 1/8 Hispanic / Latinx | 2/24 Hispanic / Latinx |
| Race | 1/16 Black, 14/16 White, 1/16 Other | 7/8 White, 1/8 Mixed race | 1/24 Black, 1/24 Mixed race, 1/6 Other race |

**Fig. 1.**
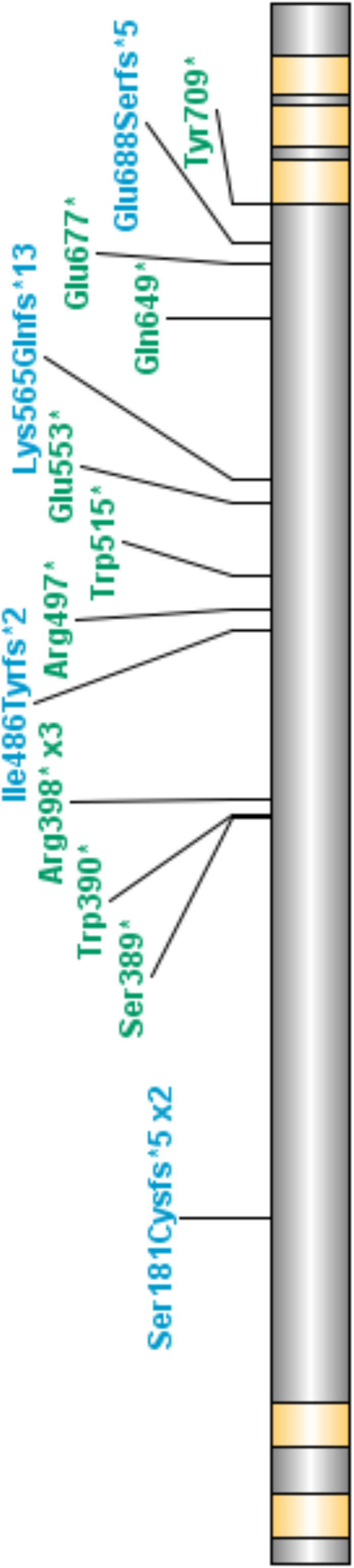
Genetic Details [place here]. a. Mutations in *CHAMP1* aligned on the protein. Blue text represents frameshift variants and green text represents nonsense variants. Orange bars represent zinc finger domains. b. Deletions of the *CHAMP1* gene. Alternating black and grey bars indicate chromosome bands, black bars represent the coordinates of participant deletions. OMIM genes are listed below participant deletions, green text indicates the gene can be disease causing.

**Fig. 2.**
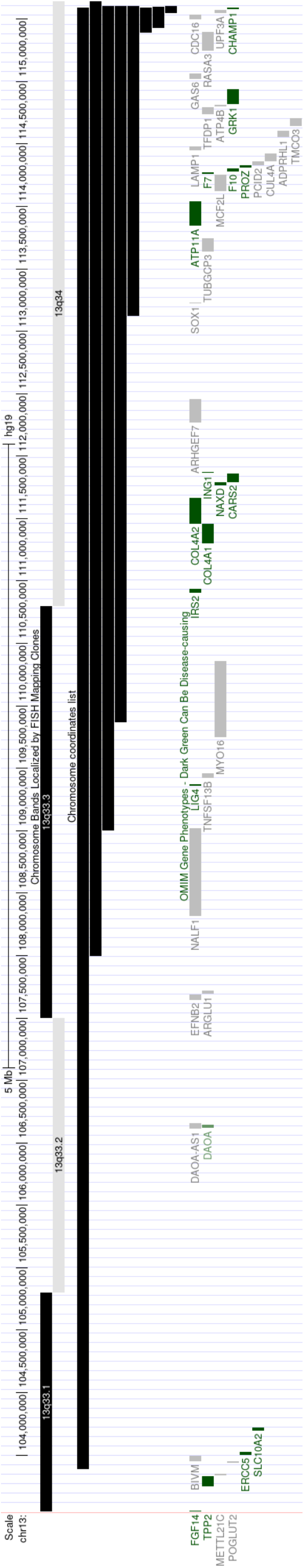
Vineland-3 Communication Domains [place here]. Boxplots representing Vineland-3 standard scores. The horizontal lines represent the minimum and maximum score within a group, the ends of the box represent the upper and lower quartiles, and the line within the box represents the median. The dark purple represents the deletion group, and the light blue represents the mutation group. Asterix represent significance, * p<=0.05, **, p<0.01, *** p<=0.001, **** p<=0.0001

**Fig. 3.**
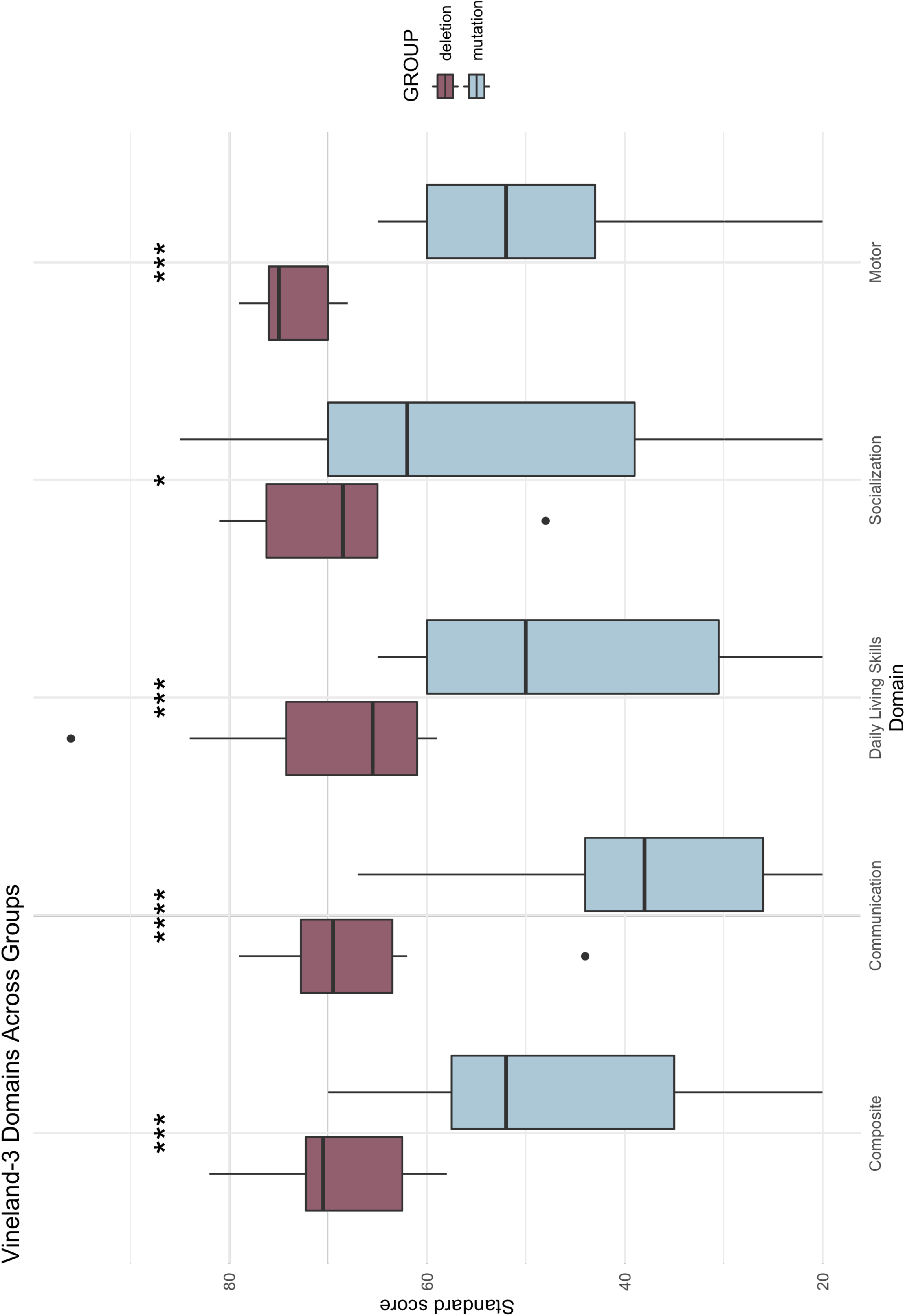
Distribution of Vineland-3 Standard Scores [place here]. Density histograms and normal curves representing the distribution of Vineland-3 domain standard scores. The light blue bars/line represent the mutation group, the dark purple bars/line represent the deletion group, and the black line represents the general population (M=100, SD=15).

### Recruitment

Participants were included from three sources: 1) participants from our previous phenotyping study completed on CHAMP1 disorder (Levy 2022b) (n=11), 2) recruitment and phenotyping completed at the CHAMP1 Family Foundation Conference 2022 (n=6), and 3) remote phenotyping completed post-conference in 2022 and 2023 (n=9, 7 included). For all sources, TL and PMS directly carried out and/or supervised the phenotyping.

Our first study on CHAMP1 disorder began during the COVID-19 pandemic; as such, we were limited to remote phenotyping. The aim of this study was to learn as much as possible about the clinical phenotype using telehealth; approximately 5 hours of participant time and 15 hours of caregiver time were given from each family to complete an extensive battery of clinical testing. During the CHAMP1 Family Foundation conference, to make the most of participants being in-person and to recruit as many participants as possible, we chose three direct testing measures totaling 1 hour of participant time and an additional 3 hours of caregiver time. Lastly, to recruit additional participants with deletions, an abbreviated remote battery was completed, consisting of 3 hours of caregiver time. As most participants with deletions were international, only remote phenotyping was feasible. Of the deletion participants, 2/8 were within the United States, 2/8 in Canada, and 1/8 in each Iceland, Australia, Belgium, United Kingdom, and Denmark.

For the current study, we compared measures that were shared across assessment batteries regardless of recruitment methods. Adaptive functioning was measured by the Vineland Adaptive Behavior Scales – Third Edition Survey Interview Form (Hill 2017). Behavior was measured using the Aberrant Behavior Checklist (ABC) (Aman 1994). Vocabulary was measured using the MacArthur Bates Communicative Index (Fenson 2007). Basic medical history and developmental milestones were assessed for all participants by clinician interview (n=11) or caregiver survey (n=13).

### Statistics

T-tests and chi-squares assessed differences between the mutation and deletion groups. Benjamini-Hochberg corrections were used to correct for multiple comparisons, which used an α level of 0.05. Rstudio was used to complete analyses and for figure design. Exploratory analyses examined correlations in the deletion group between deletion size and Vineland-3 domains and in the mutation group between amino acid location and Vineland-3 domains.

## Results

### Genetics

Within the mutation group, participants most commonly had nonsense variants (n=11) followed by frameshift variants (n=5). Deletion sizes ranged from 57.7 kilobase pairs (kb) to 11,705.7 kb. Three participants carried a separate duplication, one affecting 16q24.3, another affecting 18q23, and the third affecting Xp22.31. Sizes of these duplications were 448,465 kb, 2,541,590 kb, and 1,476,585 kb, respectively. The duplication affecting 18q23 was a confirmed translocation. Two additional participants with ring chromosomes were identified but were not included in the deletion group due to genetic differences in etiology. There were not enough participants with a ring chromosome to warrant a third group for comparison; as such, these participants are described briefly below, and descriptive data are available in Supplemental Table 1.

### Adaptive Functioning

All participants were administered the Vineland-3 Parent Interview Form. Within the Communication Domain, individuals in the deletion group had significantly higher scores in Receptive, Expressive, Written Language subdomains, and overall Communication (Table 3). The average Communication score was 2 standard deviations higher in the deletion group than the mutation group. Within the subdomains, Expressive Language was the most striking difference, with the deletion group having an average score 2.3 standard deviations higher than the mutation group.

**Table 3.** Vineland-3 Results.

| **Table 3. Vineland-3 Results** |  |  |  |  |
| --- | --- | --- | --- | --- |
| **Domain** | **Mutation** | **Deletion** | ***p*** | **Effect size** |
| **Communication** | 37.44 (15.5) | 67.12 (11.1) | 3.41E-05* | 2.20 |
| Receptive | 6.59 (3.7) | 9.88 (1.5) | 0.001* | 1.17 |
| Expressive | 2.00 (2.1) | 9.00 (4.7) | 0.003* | 1.92 |
| Written | 3.33 (3.6) | 7.71 (2.3) | 0.005* | 1.45 |
| **Daily Living Skills** | 43.50 (17.5) | 70.38 (13.1) | 0.001* | 1.74 |
| Personal | 2.25 (2.3) | 6.75 (5.5) | 0.055 | 1.07 |
| Domestic | 6.58 (2.9) | 10.29 (3.2) | 0.028* | 1.21 |
| Community | 4.25 (3.8) | 7.86 (2.1) | 0.016* | 1.18 |
| **Socialization** | 53.81 (20.0) | 68.88 (10.7) | 0.025* | 0.94 |
| Interpersonal | 6.57 (4.3) | 8.88 (3.2) | 0.187 | 0.56 |
| Play and Leisure | 5.69 (4.1) | 8.75 (4.0) | 0.102 | 0.76 |
| Coping | 7.00 (2.6) | 9.57 (2.2) | 0.028* | 1.07 |
| **Motor** | 49.67 (13.6) | 73.6 (4.5) | 0.001* | 2.36 |
| Gross Motor | 5.00 (2.7) | 10.00 (2.5) | 0.006* | 1.92 |
| Fine Motor | 6.33 (3.2) | 9.6 (2.7) | 0.072 | 1.10 |
| **Adaptive Behavior Composite** | 45.75 (15.9) | 68.5 (8.1) | 1.30E-04* | 1.80 |
| Bolded variables are domain scores, measured in standard scores, with a population mean of 100 and standard deviation of 15. Non-bolded variables are measured in V-Scale scores, with a population mean of 15 and standard deviation of 3. Some subdomains are completed for specific age groups; twelve and seven participants in the Mutation and deletion group, respectively, completed the Written, Domestic, and Community domains. Nine and five participants from the Mutation and deletion group, respectively, completed the Motor Skills domain, including both subdomains. Effect size is Cohen's D, where 0.2~small effect, 0.5~moderate effect, 0.8~ large effect. * Represents significance after correction for multiple comparisons. |  |  |  |  |

The overall Daily Living Skills Domain score was significantly higher in the deletion group compared to the mutation group, with the average score being almost 2 standard deviations higher when compared to the mutation group. The Community and Domestic subdomains were also significantly different, where the deletion group had higher scores. The Personal domain fell just below statistical significance and represents a relative weakness in both groups. The Socialization domain score showed significant differences between groups, with a difference of over 1 SD. The Motor domain and the Gross Motor subdomain showed significant differences between groups, where the deletion group scored higher than the mutation group.

### Behavior

Thirteen individuals in the mutation group and eight in the deletion group completed the ABC. Higher scores indicate a greater number of symptoms. Within the mutation group, average scores were 14.85±9.0 for Irritability, 6.31±5.9 for Social Withdrawal, 3.15±3.4 for Stereotypy, 18.77±9.1 for Hyperactivity, and 4.08±3.5 for Inappropriate Speech. In the Deletion group, average scores were 10.12±6.6 for Irritability, 3.88±2.6 for Social Withdrawal, 2.38±1.8 for Stereotypy, 12.25±6.3 for Hyperactivity, and 2.62±1.4 for Inappropriate Speech. Though the deletion group tended to have lower average scores, the domains were not significantly different between groups, with p values being 0.183, 0.214, 0.501, 0.068, and 0.205 for each domain, respectively.

### Developmental Milestones

Achievement of milestones and time of achievement was collected for all participants (Table 5). Fifteen of 16 participants in the mutation and 8/8 in the deletion group achieved sitting independently and crawling; the participant who did not yet achieve these milestones was less than 3 years old. Two participants in the mutation group and one in the deletion group had not yet walked; the two in the mutation group were both under 3 years old, and the participant in the deletion group was less than 2 years old. Thirteen of 16 participants in the mutation group and all participants in the deletion group had achieved a first word. Participants in the mutation group without a first word were all younger than the average age of achievement (1-3 years) for their group. Half (8/16) of the mutation group compared to over 85% (7/8) of the deletion group achieved two-word phrase speech. The participant in the deletion group without a phrase was under 2 years old, and the participants in the mutation group ranged from 1-16 years old, with 3 being above the average age of achievement in that group. Ages of achievement are in Table 5. Participants with deletions developed walking and their first two-word phrase significantly earlier than those with mutations.

**Table 5.** Developmental Milestones and Vocabulary.

| **Table 5. Developmental Milestones and Vocabulary** |  |  |  |  |  |
| --- | --- | --- | --- | --- | --- |
| **Milestone** | **Mutation** | **Deletion** | ***p*** | **Effect size** | **Population Average** |
| Sitting | 11 (4.5) | 8 (2.8) | 0.173 | 0.80 | 6-9 |
| Crawling | 20 (16.2) | 11 (3.8) | 0.064 | 0.76 | 7-10 |
| Walking | 27 (11.5) | 18 (5.1) | 0.017* | 1.01 | 12 |
| First Word | 35 (39.2) | 17 (6.6) | 0.129 | 0.64 | 12 |
| Two-Word Phrase | 82 (47.3) | 27 (10.4) | 0.014* | 1.61 | 24 |
| Words Understood | 248 (160.6) | 350 (116.9) | 0.095 | 0.73 | n/a |
| Words Produced<sup>1</sup> | 45 (84.9) | 309 (149.0) | 0.001* | 2.18 | n/a |
| For milestones, values represent average and standard deviation of the age in months at milestone achievement. For Words Understood and Produced, values are measured by the MacArthur Bates Communicative Index and represent number of words. Effect size is Cohen's D, where 0.2~small effect, 0.5~moderate effect, 0.8~large effect. * Represents significance after correction for multiple comparisons. |  |  |  |  |  |

Though the other milestones seem to show differences (e.g., almost double the time to develop a first word), they did not meet statistical cutoff for significance. The estimated population average for these skills is listed in Table 5 to aid in comparison to typically developing children.

Vocabulary, as measured by the MacArthur Bates Communicative Index, showed significantly higher Words Produced in the deletion group as compared to the mutation group (Table 5). While differences appear between the groups for Words Understood, the difference was not significantly different.

### Medical History

Basic medical history was taken from all participants (Table 6). Comorbid medical features were more common in the mutation group than the deletion group, with the exception of heart defects and renal/urinary tract abnormalities. Hypotonia and gastrointestinal abnormalities were the only medical features with significant differences between groups, with higher prevalence in the mutation group. Gastrointestinal abnormalities, specifically cyclical vomiting, was reported as an impactful problem for CHAMP1 individuals and families.

**Table 6.**
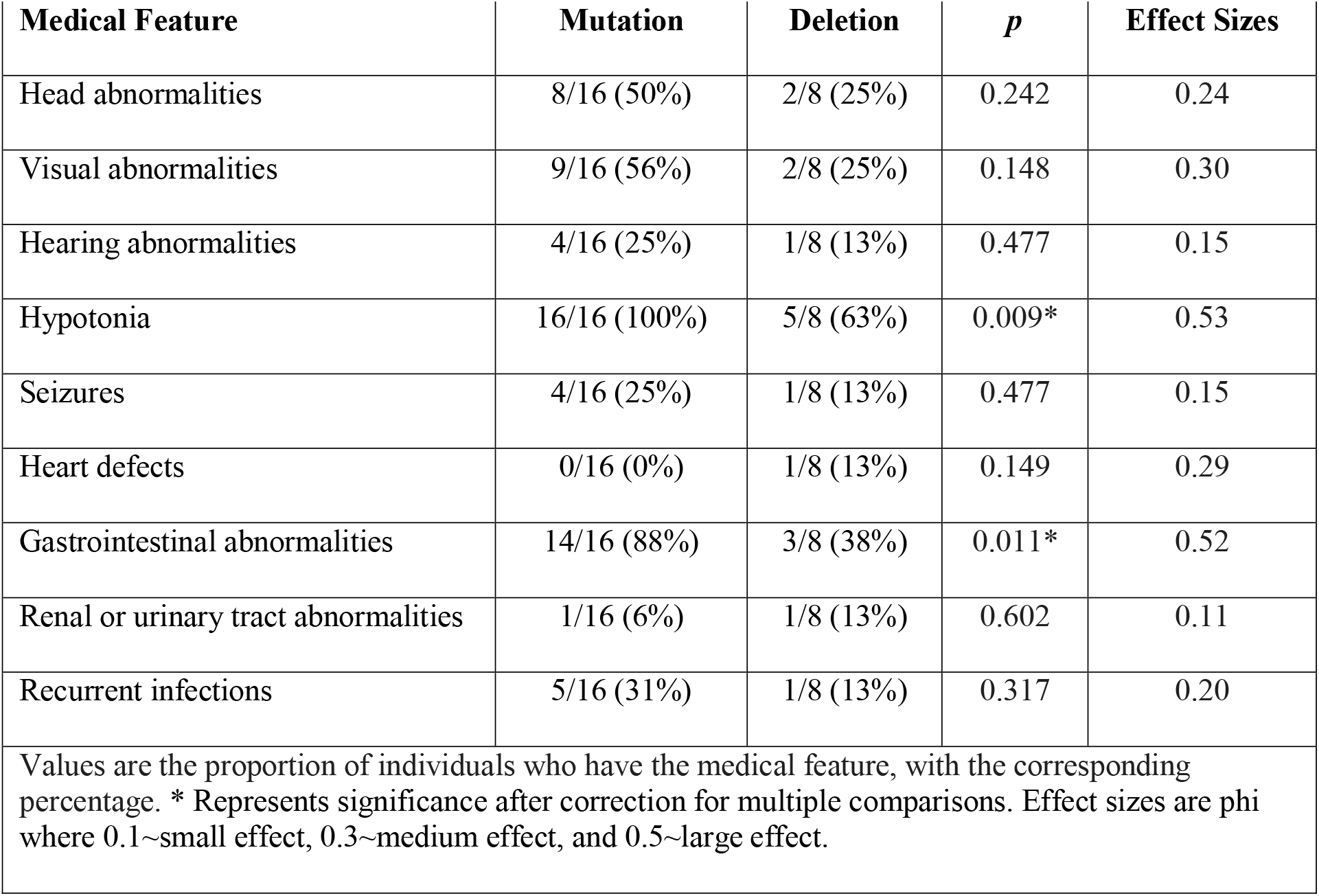
Medical Features.

| **Table 6. Medical Features** |  |  |  |  |
| --- | --- | --- | --- | --- |
| **Medical Feature** | **Mutation** | **Deletion** | ***p*** | **Effect Sizes** |
| Head abnormalities | 8/16 (50%) | 2/8 (25%) | 0.242 | 0.24 |
| Visual abnormalities | 9/16 (56%) | 2/8 (25%) | 0.148 | 0.30 |
| Hearing abnormalities | 4/16 (25%) | 1/8 (13%) | 0.477 | 0.15 |
| Hypotonia | 16/16 (100%) | 5/8 (63%) | 0.009* | 0.53 |
| Seizures | 4/16 (25%) | 1/8 (13%) | 0.477 | 0.15 |
| Heart defects | 0/16 (0%) | 1/8 (13%) | 0.149 | 0.29 |
| Gastrointestinal abnormalities | 14/16 (88%) | 3/8 (38%) | 0.011* | 0.52 |
| Renal or urinary tract abnormalities | 1/16 (6%) | 1/8 (13%) | 0.602 | 0.11 |
| Recurrent infections | 5/16 (31%) | 1/8 (13%) | 0.317 | 0.20 |
| Values are the proportion of individuals who have the medical feature, with the corresponding percentage. * Represents significance after correction for multiple comparisons. Effect sizes are phi where 0.1~small effect, 0.3~medium effect, and 0.5~large effect. |  |  |  |  |

### Ring Chromosomes

The two participants with ring chromosomes tended to look more similar to those with Mutations as compared to Deletions. On the Vineland-3, average domain standard scores were in the 20s, and V-scale scores were all 1-2, with the exception of Domestic and Coping subdomains which were relatively stronger for both participants. Behavioral, Developmental, Language, and Medical features are described in Online Resource 1.

### Exploratory analysis of deletions

Although the sample size was moderate, deletion size was not correlated with the Vineland-3 Adaptive Behavior Composite (t=-1.17, p=0.29), Communication (t=-1.73, p=0.13), Daily Living Skills (t=-1.28, p=0.25), or Socialization (t=0.06, p=0.96) domains. The Motor Domain was not assessed as only 5 participants fell within the required age range.

### Exploratory analysis of mutations

Location of amino acid change was not correlated with the Vineland-3 Adaptive Behavior Composite (t=-1.01, p=0.33), Communication (t=-0.80, p=0.44), Daily Living Skills (t=-0.75, p=0.46), Socialization (t=-1.00, p=0.34), or Motor (t=-0.62, p=0.56) domains. Visual inspection of scores plotted with amino acid position did not suggest any non-linear relationships.

## Discussion

This study provides the first direct comparison of mutation type in a cohort of prospectively characterized individuals with genetic alterations of *CHAMP1*. We characterized individuals with 13q3 deletions encompassing *CHAMP1* and additional individuals with CHAMP1 mutations. Individuals with deletions show significantly better adaptive functioning and faster developmental skill attainment compared to individuals with *CHAMP1* mutations. All domains and almost all subdomain scores on the Vineland-3 were significantly lower in individuals with genic mutations. Medical features tended to be more common in the mutation group. Socializing and relationships have been identified as relative strengths of individuals with CHAMP1 mutations (Hempel 2015; Levy 2022b), and in the current study, while there are significant differences in the Socialization domain of the Vineland-3, these differences were less pronounced compared to other domains. There was a range of ability within both groups, illustrating that while these groups are distinct from one another, neither is homogenous. Finally, though individuals with deletions did tend to have better adaptive skills, this group still displayed significantly impaired adaptive skills compared to the general population. Similarly, while individuals with deletions tended to achieve milestones earlier than individuals with mutations, milestones were still delayed in both groups. Other differences between groups showed only trend level association but high effect sizes, suggesting that many of these observations would be significant in larger cohorts.

The deletion group had less intra-group variability, despite the wide range of genetic deletion sizes. Exploratory analysis of deletion size and Vineland-3 variables did not show any significant associations. However, some values approached significance and larger sample sizes are necessary to determine possible associations. Comparing the deletion group reported here to previous literature on 13q3 deletions shows consistent findings. Although less than 25 individuals are reported in the literature to date, basic features such as developmental delay, mild to severe ID, cardiac malformations, and dysmorphic features are reported (Huang 2012; Kirchhoff 2009; McMahon 2015; Reinstein 2016; Sagi-Dain 2019; Yang 2013). In their study, Reinstein et al noted that, though *CHAMP1* is included in the deleted region, participants appeared to be more mildly affected compared to what had been reported for CHAMP1 disorder. They speculated that there may be differing mechanisms but could did not directly assess and compare individuals with deletions and mutations. Importantly, among these reports, there are examples of inherited deletions, which is generally a sign of less severe phenotypes. In contrast, PTC mutations have so far always been de novo, consistent with a more deleterious impact of de novo mutations.

Our results provide evidence that individuals with CHAMP1 mutations are more affected than those with deletions of *CHAMP1*, although both display clinically significant developmental delays and adaptive impairments. The clinical differences observed here indicate differing pathobiology of these two groups: Deletions appear to be acting through a haploinsufficiency mechanism, while mutations are likely functioning as dominant negative of GoF mechanisms. Functional studies are warranted to further dissect the mechanism of action of clinical CHAMP1 PTC mutations. A recent manuscript, published after the completion of this work, reviewed the difference between two participants, one with a deletion of *CHAMP1* and one with a mutation within the gene, finding more mild features in the participant with the deletion (Amenta 2023). The authors hypothesize that loss of function variants lead to dominant negative effects resulting in a phenotype with severe ID, autism, and dysmorphic features, while haploinsufficiency results in borderline ID. Our results and analyses are consistent with this hypothesis. For example, expression of truncated CHAMP1 (a gene which escapes NMD) would lead to protein products that might still bind to the spindle, MAD2L1 and other cellular components, but would not bind to HP1 and POGZ, and would not localize to the chromosomal appropriately. This could likely result in a dominant negative or possibly GoF function.

Ascertainment bias is a limitation of most genetic studies, where individuals offered sequencing tests (panel testing, exome sequencing) are often more severely affected than those offered microarrays, which have been the standard of care for developmental disabilities for over ten years. It may be predicted that the mutation group is more severely affected because those individuals were more likely to be offered this type of test. However, we would expect to see more severely affected individuals with deletions if both genetic etiologies caused similar features. So, while we cannot rule out all ascertainment bias, even if it were play here, we are confident that the differences observed in our cohort are not solely due to biases due to differing strategies for genetic testing. Much of our testing was remote, but we have been able to compare in-person and remote assessment in CHAMP1 disorder, both for the same (n=4) and for independent subjects, and conclude that the remote protocol is reliable for the measures presented here.

In the era of novel therapies for neurogenetic disorders, including gene therapies, it is critical to elucidate whether mutations operate through LoF, GoF, or dominant negative mechanisms. A knockout mouse would have construct validity for a LoF mechanism, but only a mouse with a knockin of a clinical mutation would have construct validity for GoF or dominant negative mechanisms. In addition, when considering gene therapies, one would want to suppress the expression of GoF or dominant negative alleles, while for LoF alleles expression of the native protein would be an optimal strategy. From the results reported here, we would argue that suppression of CHAMP1 PTC alleles would be appropriate and that this should be further examined in vitro and in model systems.

Overall, this study provides the first direct and prospective comparison of the clinical phenotype caused by mutations in *CHAMP1*, relative to deletions encompassing *CHAMP1*. Clear clinical differences were identified between groups, with the deletion group having greater skills compared to the mutation group, despite clinically significant impairments present in both groups. Future studies with larger cohorts and with in vitro and in vivo functional studies are both warranted to further classify these differences. Overexpression of native CHAMP1 as a therapeutic strategy appears to be clearly justified in the case of 13q3 deletions but should be approached with extreme caution when considering CHAMP1 mutations. More broadly, care should be taken before assuming PTC mutations are necessarily acting through LoF mechanisms.

## Data Availability

The datasets generated during the current study are available from the corresponding author on reasonable request and IRB approval.

## Acknowledgements

We would like to thank Richelle Wissink who was instrumental in the completion of this study, and Jeff and Katis D’Angelo for their consistent support through the CHAMP1 Family Foundation. We would also like to thank the families who participated in our studies for their time and effort and for sharing the stories of their wonderful children with us.

**Online Resources**

**Online Resource 1**. Results from the Vineland-3 from participants with Ring chromosome 13.

## Author Contributions

All authors contributed to the study conception and design. Material preparation, data collection and analysis were performed by Tess Levy, Thariana Pichardo, and Hailey Silver. The first draft of the manuscript was written by Tess Levy and all authors commented on previous versions of the manuscript. All authors read and approved the final manuscript.

## Ethics Approval

This study was performed in line with the principles of the Program for Protection of Human Subjects at the Icahn School of Medicine at Mount Sinai.

## Consent to Participate

Written informed consent was obtained from all participant’s parents or legal guardians in the study.

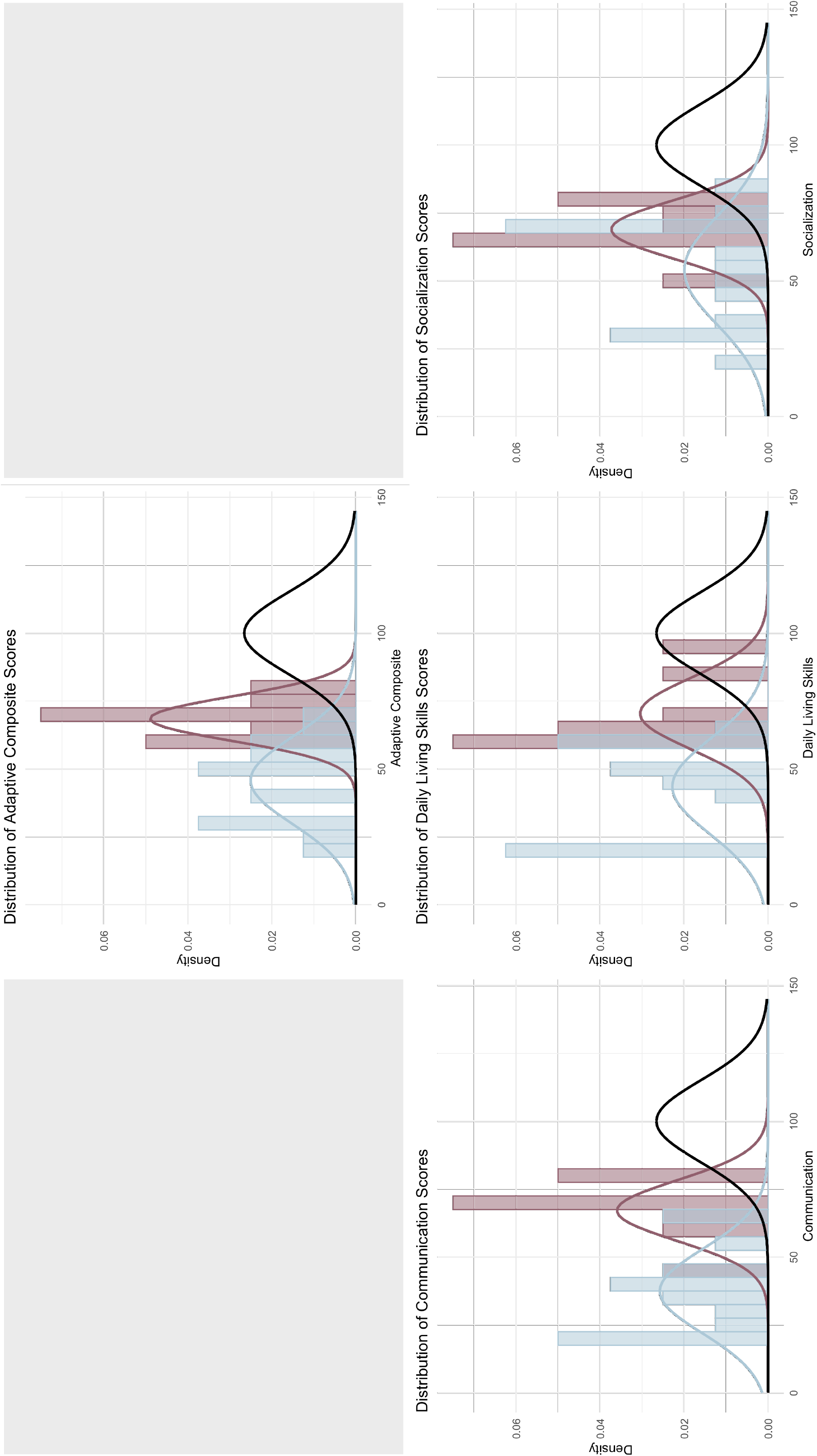

